# The health, wellbeing and health service use of women attempting or planning pregnancy: a nationwide cross-sectional survey of Australian women

**DOI:** 10.1101/2024.09.21.24314115

**Authors:** Amie Steel, Danielle Schoenaker, Joanna Harnett, Jon Adams

## Abstract

**Background:** Health and wellbeing during the preconception period influence maternal and child health outcomes. We describe the sociodemographic and health characteristics of Australian women currently attempting pregnancy or planning to conceive in the next 12 months, compared with other women of reproductive age, to identify opportunities for preconception care.

**Methods:** A sub-analysis of survey data obtained in 2022 that included 874 responses from females aged 18-49 years was conducted. Socio-demographic characteristics, health and wellbeing status, and health service utilization data were described according to pregnancy intention.

**Results:** Of 874 women, 64 (7.4%) reported currently attempting pregnancy and 45 (5.2%) reported planning to become pregnant in the next 12 months. Both groups of women were commonly married or in a de facto relationship. Women who were planning to become pregnant were more commonly aged 25-34 years (71.1%) compared to 18 to 24 years (20.0%) or aged 35 years or older (8.9%). They were also more likely to consult a chiropractor (OR 1.5). Women currently attempting pregnancy were less likely to not be in the paid workforce (OR 0.34) compared to working full time. They were also less likely to be using prescription-only pharmaceuticals (OR 0.30) and more likely to be consulting a Traditional Chinese medicine practitioner (OR 2.66) or a dietician (OR 2.11).

**Conclusion:** The findings of this study can be used for informing health service planning and policy that takes a whole-of-health-system approach when designing preconception health primary care interventions.

## Introduction

The time before conception is a critical period for preventive health providing an opportunity to improve the physical and mental health and wellbeing of both reproductive parents and their future child(ren).^1^ A range of behavioural, medical and social risk factors - including low folate intake, diet quality, smoking, alcohol and illicit drug use, obesity, poorly controlled diabetes and teratogenic medication use - are known to have important detrimental impacts on pregnancy, maternal and child health outcomes.^2^ Preconception health of both parents can be optimised, and risks reduced, through a range of interventions targeting all people of reproductive age, or those who are planning pregnancy.^3-6^ For example, folic acid fortification of flour and cereals is mandatory in over 80 countries to reduce rates of neural tube defects,^7^ and national and international preconception care guidelines encourage healthcare professionals to support their patients to plan and prepare for pregnancy.^8^

The opportunity to improve an individual’s health prior to pregnancy is linked to their level of pregnancy intention, their knowledge and capability to proactively optimise their health and their identification and support based on routine pregnancy intention and risk factor screening by health and social care professionals. In Australia,^9^ and internationally,^10-12^ at least three out of four pregnancies are estimated to involve some level of planning.^9,12,13^ Despite this, there are low levels of awareness among people of reproductive age about the importance of parental health in the preconception period and the actions they could take to prepare for a healthy pregnancy.^14-16^ As a result, poor preconception health behaviours remain common and changes in preconception health behaviours with increasing pregnancy intention modest, even among those actively planning pregnancy.^9,12,17-19^

Health professionals have a pivotal role in identifying couples who are intending to conceive and in supporting them to optimise their health behaviours and the management of physical and mental health conditions, to reduce risks in preparation for pregnancy. Primary care professionals, such as general practitioners (GPs), practice nurses and pharmacists, are most often identified as well placed to provide such preconception care,^6,14,20^ while community support workers, sexual health professionals and specialist doctors also have an important role.^3,14^ It is important for effective preconception workforce planning to identify the preconception needs of people of reproductive age, in particular among those who are planning or attempting pregnancy. While previous research has largely focused on health behaviours,^9,12,18,19^ little is known about the wellbeing, physical and mental health conditions, and product and treatment use of people planning or attempting pregnancy, and the health professionals they regularly consult.

This study therefore aimed to describe the self-reported health status, health-related quality of life and health service use of a nationwide sample of Australian women currently attempting pregnancy or planning to conceive in the next 12 months, compared with other women of reproductive age.

## Materials and Methods

### Study design and setting

This study presents a sub-analysis of data collected though cross-sectional survey research conducted in Australia. The primary study sampled adults in Australia (aged 18 years and over) to investigate use and experiences of a range of health services and products. A sub-sample of this primary study was used for this analysis.

### Participants

The primary dataset included eligible responses from adults residing in Australia (n=2561) recruited through purposive convenience sampling of a database of people who were registered with a research company to participate in research. Recruitment and data collection were conducted between 4 February and 18 February 2022. Respondents who completed the survey received a small financial incentive based on the time taken to complete the survey. Informed consent was obtained once respondents had read the information page presented prior to beginning the survey. Data were screened for disengaged and missing responses (identified by discrepancies between responses, text responses incongruous with the corresponding question, lack of variance, and repeated patterns in the data).

The primary dataset was then limited to only include respondents who identified as female, aged between 18 and 49 years (within the definition of women of reproductive age [15-49 years] by the World Health Organisation^21^) and not currently pregnant.

### Survey instrument

The survey instrument used to collect the primary data includes 63 items, of which 15 may repeat up to five times, depending on the number of responses provided to previous answers (number of health condition diagnoses, number of health professionals consulted). These questions span five domains: demographics (11 items), health and wellbeing status (20 items), health product and service use (16 items), patient experiences of care (10 items), and disclosure of medicines (6 items). This analysis employed items from the first three domains.

#### Demographics

Participants were presented with 11-items with categorical response options covering demographic characteristics including gender, age range, financial manageability, highest educational qualification, private health insurance cover, Health Care Card cover, employment status and relationship status. These were all presented at the end of the survey, with a second item about participant age also presented as a screening item at the beginning of the survey using a conditional open-text response option that only permitted data entry of whole numbers.

#### Health and Wellbeing Status

The Health and Wellbeing Status domain consisted of two validated instruments and additional items examining participant health history. The first validated instrument was the Personal Wellbeing Index (PWI)^22^ consisting of nine items designed to measure subjective wellbeing inclusive of standard of living, personal health, achieving in life, personal relationships, personal safety, community-connectedness, future security, spirituality, and life as a whole. In 2021, the normative range for the PWI among women in the Australian population was between 70 and 80.^23^ The second instrument used was Short Form-20 (SF-20) to measure health-related quality of life (HR-QOL). This well-established instrument includes 20 items covering six domains: physical functioning, role functioning, social functioning, mental health, health perceptions, and pain.

Health information was also collected via an item inviting participants to select any of 35 response options to indicate any chronic health condition(s) they had been diagnosed with or treated for in the previous three years. Alternatively, they could select ‘none of the above’. Respondents were also invited to identify if they (or their partner) were currently pregnant.

Those who indicated they were not pregnant were then presented with items to identify if they were currently attempting to become pregnant (‘Are you or your partner currently attempting to become pregnant?’: yes/no/unsure) or planning to become pregnant in the next 12 months (‘Do you or your partner plan to become pregnant in the next 12 months?’: yes/no/unsure).

#### Health product and service use

Participants were asked to provide information about their use of a range of 12 health products or treatments in the previous 12 months inclusive of prescription and over-the-counter pharmaceuticals, herbal medicines, vitamin or mineral supplements, and yoga practice. They also had the option to self-report ‘other’ products or treatments they may have used. Respondents were also invited to report the frequency of visits (5-point scale: ‘None’ to ‘More than 6’) to 17 types of health professionals including medical doctors (e.g., general practitioner, specialist doctor), registered allied health professionals (e.g., pharmacist, osteopath, acupuncturist) and other types of non-registered health professionals (e.g., dietician, naturopath, yoga teacher).

### Data management

Data were initially cleaned to recode items to optimize the analysis for statistical power. This includes regrouping the relationship status (e.g., combining ‘married’ and ‘defacto’), highest qualifications (e.g., combining ‘trade/apprenticeship’ and ‘certificate/diploma’), and financial manageability (e.g., combining ‘it is impossible’ and ‘it is difficult all the time’) items. New binary (yes/no) variables were also generated from the items reporting frequency of visits to each type of health professional. Variables were also generated to count the total number of health products or treatments, and total number of health services used.

In accordance with the PWI scoring guide, mean scores for each PWI item and the Overall PWI were calculated.^22^ Following this, individual responses where the Overall PWI was 10 or less, or 100 were converted to missing (n=10) for the PWI mean scores.^22^ Mean SF-20 Domain scores were also determined in alignment with the SF-20 scoring manual.^24^ These were then adjusted to a scale of 100 to enable comparison across domains.

A new case classification variable was also generated that grouped respondents into one of four categories: (1) not attempting pregnancy or planning pregnancy in the next 12 months, (2) currently attempting pregnancy, (3) planning to become pregnant in the next 12 months but not currently attempting, (4) unsure about attempting or planning pregnancy.

### Data analysis

Socio-demographic characteristics, health and wellbeing status, and health service utilization were described using frequencies and percentages (categorical variables) and medians with interquartile ranges (continuous variables) according to pregnancy intention. Tests of association were conducted using chi-square tests (for binary categorical variables) and analysis of variance analysis (ANOVA) (categorical and continuous variables).

To explore if these initial tests of association may be influenced by confounding, multinomial logistic regression was applied to the data, defined by the respondents self-reported pregnancy intention (case classification) with ‘not attempting pregnancy or planning pregnancy in the next 12 months’ as the baseline comparison group. The potential predictors for the regression model were selected if they were found to have an α value equal to or less than 0.1 resulting from the tests of association. Only variables that were determined through the multinomial regression model to be statistically significant, defined as an α value equal to or less than 0.05, were reported in the results.

### Patient and Public Involvement

## Results

Responses from 871 women of reproductive age were included in the analysis, of whom 745 (85.7%) reported not attempting pregnancy or planning to become pregnant in the next 12 months, 64 (7.4%) reported currently attempting pregnancy, 45 (5.2%) reported planning to become pregnant in the next 12 months but not currently attempting, and 16 (1.8%) selected ‘unsure’ about both currently attempting pregnancy and planning to become pregnant in the next 12 months. There was a statistically significant association between participants self-reported pregnancy intention and their age (p<0.001), relationship status (p<0.001), highest qualification (p<0.001) and possession of a Health Care Card (p=0.03) (see Table 1).

**Table 1:** Participant characteristics.

| Demographic | All non-pregnant women (18-49 years) (n=871) | Not attempting or planning pregnancy (n=745) | Currently attempting pregnancy (n=64) | Planning to become pregnant in next 12 months but not currently attempting (n=45) | Unsure about attempting or planning pregnancy (n=16) | *p* |
| --- | --- | --- | --- | --- | --- | --- |
| Age | n (%) | n (%) | n (%) | n (%) | n (%) |  |
| 18-24 years | 240 (27.6) | 213 (28.6) | 12 (18.8) | 9 (20.0) | 6 (37.5) | <0.001 |
| 25-34 years | 290 (33.3) | 221 (29.7) | 30 (46.9) | 32 (71.1) | 7 (43.8) |  |
| 35-49 years | 340 (39.1) | 311 (41.7) | 22 (34.4) | 4 (8.9) | 3 (18.8) |  |
| Relationship status |  |  |  |  |  |  |
| Never married | 392 (45.0) | 362 (48.5) | 12 (18.6) | 13 (28.9) | 5 (31.3) | <0.001 |
| Married or De facto relationship (opposite sex) | 396 (45.5) | 311 (41.7) | 45 (70.3) | 29 (64.4) | 11 (68.8) |  |
| Married or De facto relationship (same sex) | 34 (3.9) | 30 (4.0) | 3 (4.7) | 1 (2.2) | 0 (0.0) |  |
| Separated, Divorced or Widowed | 49 (5.6) | 43 (5.8) | 4 (6.3) | 2 (4.4) | 0 (0.0) |  |
| Highest qualification |  |  |  |  |  |  |
| Up to Year 10 or equivalent | 85 (9.8) | 71 (9.5) | 7 (10.9) | 0 (0.0) | 7 (43.8) | <0.001 |
| Year 12 or equivalent | 217 (24.9) | 191 (25.6) | 14 (21.9) | 9 (20.0) | 3 (18.8) |  |
| Trade/Apprenticeship/Certificate/Diploma | 250 (28.7) | 208 (27.9) | 20 (31.3) | 21 (46.7) | 1 (6.3) |  |
| University qualification | 319 (36.6) | 276 (37.0) | 23 (35.9) | 15 (33.3) | 5 (31.3) |  |
| Employment status |  |  |  |  |  |  |
| Full time work | 325 (37.3) | 269 (36.1) | 32 (50.0) | 20 (44.4) | 4 (25.0) | 0.09 |
| Part time work | 205 (23.5) | 170 (22.8) | 13 (20.3) | 16 (35.6) | 6 (37.5) |  |
| Casual or temporary work with irregular hours | 76 (8.7) | 67 (9.0) | 6 (9.4) | 3 (6.7) | 0 (0.0) |  |
| Looking for work | 97 (11.1) | 88 (11.8) | 4 (6.3) | 2 (4.4) | 3 (18.8) |  |
| Not in the paid workforce | 168 (19.3) | 152 (20.4) | 9 (14.1) | 4 (8.9) | 3 (18.8) |  |
| Financial manageability |  |  |  |  |  |  |
| It is impossible/It is difficult all the time | 186 (21.4) | 160 (21.5) | 14 (21.9) | 8 (17.8) | 4 (25.0) | 0.7 |
| It is difficult some of the time | 300 (34.4) | 250 (33.5) | 23 (35.9) | 20 (44.4) | 7 (43.8) |  |
| It is not too bad/It is easy | 385 (44.2) | 336 (45.0) | 27 (42.2) | 17 (37.8) | 5 (31.3) |  |
| Health Care Card | 423 (48.6) | 352 (47.2) | 39 (60.9) | 20 (44.4) | 12 (75.0) | 0.03 |
| Private health insurance cover | 379 (43.5) | 322 (43.2) | 32 (50.0) | 20 (44.4) | 5 (31.3) | 0.5 |

### Health status, health-related quality of life and personal wellbeing

The median overall PWI for the full sample was 65 (Q1 53, Q3 75) which is lower than the norm for Australian women in 2021 (70 to 80) (see Table 2). The lowest domain median score was for satisfaction with your spirituality or religion (50; Q1 70, Q3 90). The PWI scores did not differ based on participant pregnancy intention. Respondents’ health-related quality of life reflected higher levels of physical (M 92), role (M 100) and social (M 80) functioning but lower scores of pain (M 20), health perceptions (M 50), and mental health (M 56). The most common type of health condition reported by participants was a mental health, psychiatric or neurological condition (43.3%). There was a statistically significant difference in health-related quality of life based on pregnancy intention across the domains of physical function (p=0.03), mental health (p=0.01) and health perception (p=0.001). No differences in diagnoses or personal wellbeing were identified as statistically significant.

**Table 2:** Health diagnoses, health-related quality of life and personal wellbeing among women currently attempting pregnancy or planning pregnancy in the next 12 months, compared to other women.

| Health diagnoses, health-related quality of life and personal wellbeing | All non-pregnant women (18-49 years) (n=858) | Not attempting or planning pregnancy (n=726) | Currently attempting pregnancy (n=63) | Planning to become pregnant in next 12 months but not currently attempting (n=45) | Unsure about attempting or planning pregnancy (n=16) | *p* |
| --- | --- | --- | --- | --- | --- | --- |
|  | *Median (Q1, Q3)* | *Median (Q1, Q3)* | *Median (Q1, Q3)* | *Median (Q1, Q3)* | *Median (Q1, Q3)* |  |
| **Personal Wellbeing Index** |  |  |  |  |  |  |
| *Satisfaction with your life as a whole (n=858)* | 70 (50, 80) | 70 (50, 80) | 70 (50, 80) | 70 (50, 70) | 60 (50, 80) | 0.7 |
| *Satisfaction with your standard of living* | 70 (50, 80) | 70 (50, 80) | 70 (50, 80) | 70 (50, 90) | 55 (40, 75) | 0.7 |
| *Satisfaction with your health* | 60 (50, 80) | 60 (50, 80) | 60 (50, 80) | 70 (50, 80) | 60 (40, 70) | 0.7 |
| *Satisfaction with what you are achieving in life* | 60 (40, 80) | 60 (40, 80) | 70 (40, 80) | 70 (50, 80) | 60 (40, 75) | 0.4 |
| *Satisfaction with your personal relationships* | 70 (50, 80) | 70 (50, 80) | 70 (50, 90) | 80 (50, 90) | 70 (45, 90) | 0.5 |
| *Satisfaction with how safe you feel* | 70 (60, 90) | 70 (60, 90) | 70 (50, 90) | 80 (70, 90) | 65 (50, 90) | 0.6 |
| *Satisfaction with feeling part of your community* | 60 (40, 70) | 60 (40, 70) | 60 (40, 80) | 50 (40, 80) | 50 (45, 80) | 0.9 |
| *Satisfaction with your future security* | 60 (40, 80) | 60 (40, 80) | 60 (40, 70) | 60 (40, 70) | 60 (45, 60) | 1.0 |
| *Satisfaction with your spirituality or religion* | 50 (70, 90) | 50 (70, 90) | 50 (70, 90) | 50 (70, 90) | 55 (70, 100) | 0.9 |
| **Overall Personal Wellbeing Index** | 65 (53, 75) | 65 (51, 75) | 69 (51, 76) | 69 (56, 75) | 61 (56, 70) | 0.9 |
| **Short Form-20 Domains** |  |  |  |  |  |  |
| *Physical function* | 92 (58, 100) | 92 (67, 100) | 83 (50, 100) | 83 (67, 100) | 83 (46, 96) | 0.03 |
| *Role function* | 100 (50, 100) | 100 (50, 100) | 100 (38, 100) | 100 (75, 100) | 75 (50, 100) | 0.07 |
| *Social function* | 80 (40, 100) | 80 (60, 100) | 80 (40, 100) | 80 (40, 100) | 50 (30, 90) | 0.08 |
| *Mental health* | 56 (48, 64) | 60 (48, 64) | 52 (40, 60) | 56 (48, 64) | 52 (40, 62) | 0.01 |
| *Health perception* | 50 (45, 55) | 50 (45, 55) | 45 (40, 50) | 50 (45, 60) | 55 (45, 60) | 0.001 |
| *Bodily pain* | 20 (20, 40) | 20 (20, 40) | 20 (20, 40) | 20 (20, 40) | 40 (20, 40) | 0.2 |
| **Diagnosed health conditions** | ***N (%)*** | ***N (%)*** | ***N (%)*** | ***N (%)*** | ***N (%)*** |  |
| *Mental health, psychiatric and neurological conditions* | 377 (43.3) | 318 (42.6) | 27 (42.2) | 21 (46.7) | 11 (68.8) | 0.2 |
| *Respiratory and immune conditions* | 218 (25.0) | 184 (24.7) | 14 (21.9) | 15 (33.3) | 5 (31.3) | 0.5 |
| *Reproductive and genitourinary conditions* | 105 (12.1) | 83 (11.1) | 10 (15.6) | 9 (20.0) | 3 (18.8) | 0.2 |
| *Cardiometabolic and endocrine conditions* | 88 (10.1) | 70 (9.4) | 11 (17.2) | 7 (15.6) | 0 (0.0) | 0.07 |
| *Other chronic conditions* | 188 (21.6) | 164 (22.0) | 15 (23.4) | 5 (11.1) | 4 (25.0) | 0.36 |

### Health product and service use

Participants reported using a range of health products including over-the-counter (56.7%) and prescription-only (54.1%) pharmaceuticals, vitamin and mineral supplements (44.9%), and relaxation techniques including mediation and mindfulness (31.5%). Least commonly used were Tai chi or Qi gong practice (1.7%), ingested aromatherapy oils (2.3%) and homeopathic remedies (3.6%). Overall, participants reported using a median of two health products or treatments in the previous 12 months. When compared across categories of pregnancy intention, a statistically significant association was identified for use of prescription-only pharmaceuticals (p=0.01) and use of homeopathic remedies (p=0.002) in the previous 12 months.

The majority of participants reported consulting with a general practitioner (87.1%) or pharmacist (70.8%) in the previous 12 months. A lower rate of visits was reported for other health professionals, with the most common being a specialist (31.1%) or hospital (21.9%) doctor, a counsellor or mental health worker (27.1%), a massage therapist (16.8%), a physiotherapist (15.2%), a community nurse or nurse practitioner (11.7%), or a yoga therapist (11.7%). There was a significant difference between participants when comparing pregnancy intention with visits to all health professionals *except* general practitioner, pharmacist, specialist doctor, physiotherapist and osteopath. The total sample of respondents reported visiting a median of three types of health professionals in the previous 12 months, although this varied significantly when compared across pregnancy intention categories (p<0.001)

### Characteristics likely associated with preconception intention

Table 4 presents the results of the multinomial regression. Survey participants who identified as ‘unsure’ about currently attempting pregnancy or planning to become pregnant in the next 12 months were excluded from the analysis due to the small number of participants in that group. Compared with participants who were not currently attempting or planning pregnancy, those currently attempting pregnancy were more likely to be married or in a *de facto* relationship with an opposite sex partner (OR 4.55, 95% CI 2.17–9.56) rather than never married, and less likely to not be in the paid workforce (OR 0.34, 95% CI 0.13–0.88) rather than in full time work, or to possess a health care card (OR 0.42, 95% CI 0.21-0.1). Participants who were currently attempting pregnancy were less likely to have used prescription-only pharmaceuticals (OR 0.30, 95% CI 0.16-0.58) and were more likely to have consulted a traditional Chinese medicine practitioner (OR 2.66, 95% CI 1.27-5.55) or a dietician (OR 2.11, 95% CI 1.11-4.01) in the previous 12 months, when compared with participants who were not attempting pregnancy or planning to become pregnant in the next 12 months.

**Table 3:** Health product used and health professionals visited in the previous 12 months.

| Health product or service | All non-pregnant women (18-49 years) (n=871) | Not attempting or planning pregnancy (n=746) | Currently attempting pregnancy (n=64) | Planning to become pregnant in next 12 months but not currently attempting (n=45) | Unsure about attempting or planning pregnancy (n=16) | *p* |
| --- | --- | --- | --- | --- | --- | --- |
|  | *n (%)* | *n (%)* | *n (%)* | *n (%)* | *n (%)* |  |
| **Product or treatments** |  |  |  |  |  |  |
| *Over-the-counter pharmaceuticals* | 494 (56.7) | 426 (57.1) | 35 (54.7) | 26 (57.8) | 7 (43.8) | 0.7 |
| *Prescription-only pharmaceuticals* | 471 (54.1) | 419 (56.2) | 23 (35.9) | 22 (48.9) | 7 (43.8) | 0.01 |
| *Vitamin/mineral supplements* | 391 (44.9) | 330 (44.2) | 33 (51.6) | 24 (53.3) | 4 (25.0) | 0.2 |
| *Relaxation techniques/meditation/mindfulness* | 274 (31.5) | 231 (31.0) | 25 (39.1) | 14 (31.1) | 4 (25.0) | 0.6 |
| *Yoga practice* | 144 (16.5) | 124 (16.6) | 8 (12.5) | 10 (22.2) | 2 (12.5) | 0.6 |
| *Aromatherapy oils (externally applied)* | 119 (13.7) | 102 (13.7) | 9 (14.1) | 7 (15.6) | 1 (6.3) | 0.8 |
| *Chinese herbal medicines* | 55 (6.3) | 47 (6.3) | 6 (9.4) | 1 (2.2) | 1 (6.3) | 0.5 |
| *Western herbal medicines* | 41 (4.7) | 34 (4.6) | 3 (4.7) | 4 (8.9) | 0 (0.0) | 0.5 |
| *Flower essences* | 40 (4.6) | 35 (4.7) | 4 (6.3) | 1 (2.2) | 0 (0.0) | 0.6 |
| *Homeopathic remedies* | 32 (3.7) | 21 (2.8) | 7 (10.9) | 4 (8.9) | 0 (0.0) | 0.002 |
| *Aromatherapy oils (ingested)* | 19 (2.2) | 14 (1.9) | 4 (6.3) | 1 (2.2) | 0 (0.0) | 0.1 |
| *Tai chi or Qi gong practice* | 15 (1.7) | 12 (1.6) | 2 (3.1) | 1 (2.2) | 0 (0.0) | 0.8 |
|  | **Median (Q1, Q3)** | **Median (Q1, Q3)** | **Median (Q1, Q3)** | **Median (Q1, Q3)** | **Median (Q1, Q3)** |  |
| Total number of health product or treatment categories used | 2 (1, 3) | 2 (1, 3) | 2 (1, 3) | 2 (1, 4) | 1.5 (1, 2) | 0.2 |
| **Health professionals consulted** | ***n (%)*** | ***n (%)*** | ***n (%)*** | ***n (%)*** | ***n (%)*** |  |
| *General practitioner* | 759 (87.1) | 648 (86.9) | 56 (87.5) | 39 (86.7) | 16 (100) | 0.5 |
| *Pharmacist* | 617 (70.8) | 519 (69.6) | 51 (79.7) | 35 (77.8) | 12 (75.0) | 0.2 |
| *Specialist doctor* | 271 (31.1) | 224 (30.0) | 26 (40.6) | 17 (37.8) | 4 (25.0) | 0.2 |
| *Counsellor or mental health worker* | 236 (27.1) | 193 (25.9) | 27 (42.2) | 15 (33.3) | 1 (6.3) | 0.006 |
| *Hospital doctor* | 191 (21.9) | 148 (19.8) | 25 (39.1) | 14 (31.1) | 4 (25.0) | 0.002 |
| *Massage therapist* | 146 (16.8) | 116 (15.6) | 18 (28.1) | 11 (24.4) | 1 (6.3) | 0.02 |
| *Physiotherapist* | 132 (15.2) | 111 (14.9) | 10 (15.6) | 11 (24.4) | 0 (0.0) | 0.1 |
| *Community nurse or nurse practitioner* | 102 (11.7) | 78 (10.5) | 16 (25.0) | 7 (15.6) | 1 (6.3) | 0.004 |
| *Yoga practitioner/instructor* | 102 (11.7) | 78 (10.5) | 11 (17.2) | 10 (22.2) | 3 (18.8) | 0.04 |
| *Chiropractor* | 81 (9.3) | 60 (8.0) | 6 (9.4) | 14 (31.1) | 1 (6.3) | <0.001 |
| *Dietician* | 68 (7.8) | 43 (5.8) | 13 (20.3) | 10 (22.2) | 2 (12.5) | <0.001 |
| *Naturopath* | 52 (6.0) | 35 (4.7) | 9 (14.1) | 6 (13.3) | 2 (12.5) | 0.002 |
| *Acupuncturist* | 49 (5.6) | 35 (4.7) | 9 (14.1) | 3 (6.7) | 2 (12.5) | 0.01 |
| *Osteopath* | 47 (5.4) | 35 (4.7) | 7 (10.8) | 4 (8.9) | 1 (6.3) | 0.1 |
| *Traditional Chinese medicine practitioner* | 45 (5.2) | 27 (3.6) | 13 (20.3) | 4 (8.9) | 1 (6.3) | <0.001 |
| *Western herbalist* | 40 (4.6) | 24 (3.2) | 11 (17.2) | 4 (8.9) | 1 (6.3) | <0.001 |
| *Homeopath* | 30 (3.4) | 21 (2.8) | 4 (6.3) | 5 (11.1) | 0 (0.0) | 0.01 |
|  | **Median (Q1, Q3)** | **Median (Q1, Q3)** | **Median (Q1, Q3)** | **Median (Q1, Q3)** | **Median (Q1, Q3)** |  |
| Total number of types of health professionals visited | 3 (2, 4) | 3 (2, 4) | 4 (2, 6) | 4 (2, 6) | 3 (2, 4) | <0.001 |

**Table 4:**
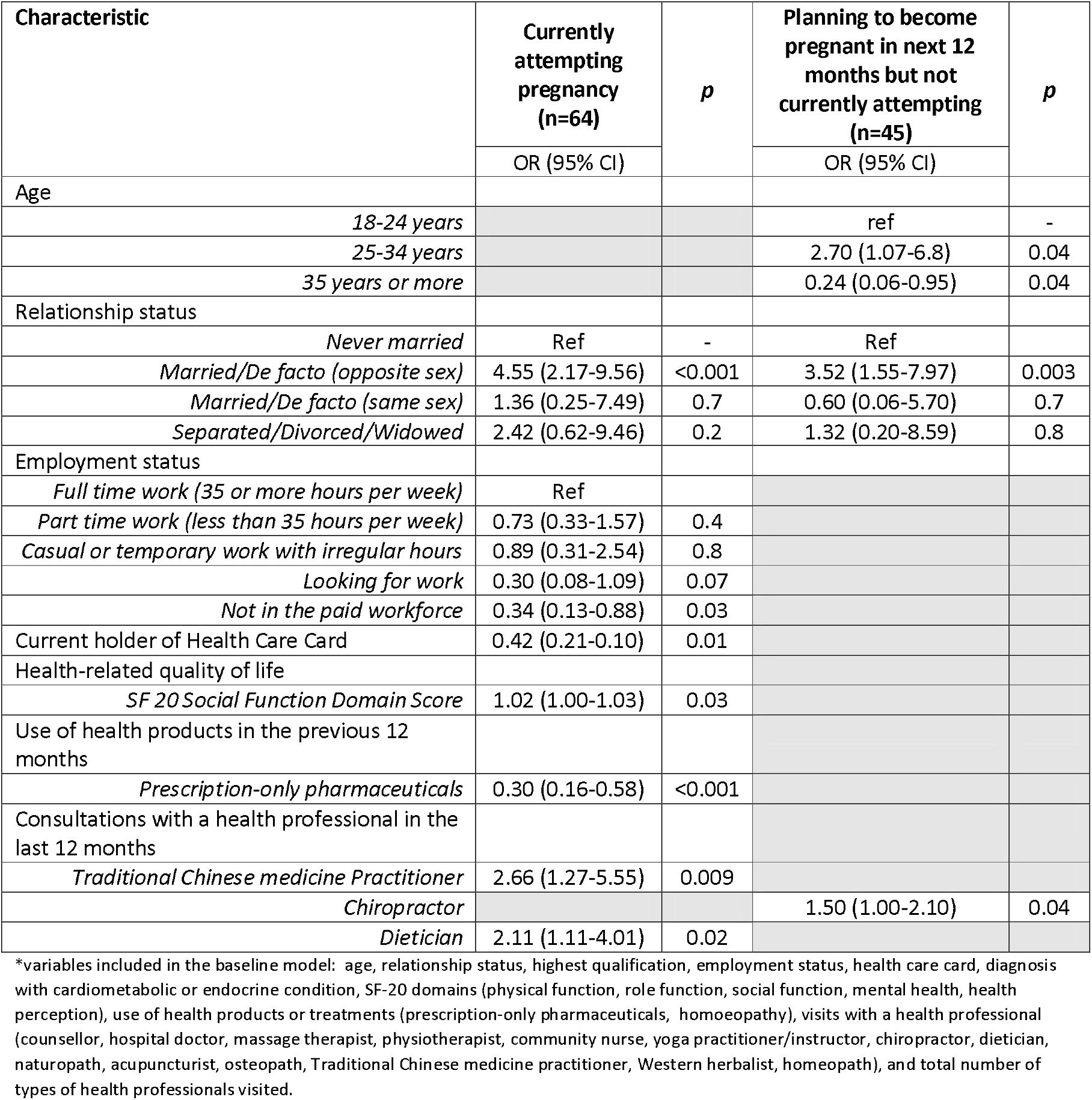
Characteristics associated with women identifying as ‘currently attempting pregnancy’ or ‘planning to become pregnant in the next 12 months but not currently attempting’, compared to ‘not attempting or planning pregnancy’ (n=???)

Study participants who reported planning to become pregnant in the next 12 months (but not currently attempting) were more likely to be 25-34 years (OR 2.70, 95% CI 1.07-6.80) and less likely to be 35 years or more (OR 0.24, 95% CI 0.06-0.95) rather than 18 to 24 years, compared to participants who were not attempting or planning a pregnancy. They were also more commonly married or in a **de facto** relationship with an opposite sex partner (OR 3.52, 95% CI 1.55-7.97) rather than never married, compared to those not attempting or planning pregnancy, and more likely to have visited a chiropractor in the previous 12 months (OR 1.5, 95% CI 1.0-2.1).

## Discussion

This nationwide study of the self-reported health status, health-related quality of life and health service use of Australian women currently attempting pregnancy or planning to conceive in the next 12 months identified a number of findings that warrant further discussion. First, a diverse range of health professionals were consulted by the sample in the previous 12 months. GPs were the most common of these health professionals, but pharmacists, other medical doctors (specialists, hospital doctors), counsellors and mental health workers, massage therapists, chiropractors, physiotherapists, were also consulted by more than one quarter of women planning or attempting pregnancy. These women visited an average of four (and commonly up to six) types of health professionals in the previous 12 months. While previous research lists GPs, practice nurses and pharmacists as the health professions most commonly identified as well placed to provide preconception care,^6,14,20^ our study finding suggests health policy and service planning needs to consider a whole-of-health-system approach when designing preconception health primary care interventions. Such an approach should be co-designed with patients, health professionals and service providers, and consider any health profession with whom an individual may engage with as first contact in the health system as primary care.^25^ Much preconception care relies on screening for modifiable risk factors and educating women and their partners about health behaviour change necessary to increase the likelihood for a health pregnancy and child.^5^ Given the low levels of awareness with regards to preconception health risk factors in the community,^14-16^ co-designed preconception health promotion interventions are urgently needed and these interventions should leverage each health care encounter through an “every contact is an opportunity” approach ^26^. With this in mind, health professionals from all areas of primary care – defined as any health profession an individual may engage with as first contact in the health system^25^ - require access to the necessary tools and training to identify, educate and facilitate health behaviour change for optimal preconception health.^27^

The analysis found women planning or attempting pregnancy had an increased likelihood of consulting with three health professions in particular: Traditional Chinese medicine practitioners, dieticians, and chiropractors. Research supports the potential contribution of these first two health professions in the preconception period. Poor diet and nutritional status, for example, is a known modifiable preconception risk factor linked with infertility, pre-eclampsia, pre-term birth, and small-for-gestational-age and other adverse outcomes.^2^ Similarly, preconception and interconception overweight and obesity are known risks for outcomes such as pre-eclampsia, stillbirth, infant macrosomia, congenital heart defects, childhood cognitive development, and childhood overweight.^2^ Dieticians are well-placed to support women minimise these health risks prior to pregnancy.^28^ Previous research has also found Traditional Chinese medicine practitioners are commonly accessed by women attempting pregnancy if they have a history of fertility issues,^29^ which aligns with clinical research suggesting acupuncture may assist with pregnancy outcomes for women undergoing IVF.^30^ Combined, this previous research may explain why consultations with a Traditional Chinese medicine practitioner or dietician are more common among women currently attempting a pregnancy rather than those planning a pregnancy in the next 12 months. In contrast, there is limited existing research to help explain why women are more likely to consult with a chiropractor when planning to become pregnant in the next 12 months. Previous research has found women consult with a chiropractor during pregnancy, primarily for musculoskeletal complaints,^31^ so it is possible that women are acting preventively in an attempt to minimise the impacts of such conditions when they become pregnant. There is also evidence that chiropractors^32,33^ and Traditional Chinese medicine practitioners^34,35^ discuss some preventive health topics with their patients such as physical activity, diet and other health behaviour changes. However, further research is needed to understand the nature of the support many health professionals provide to women during the preconception period, including the degree to which they actively enquire about pregnancy intention, their confidence in providing preconception health advice and the role they see themselves or others in their profession playing in the wider preconception care landscape.

This study also provides some insights into sociodemographic characteristics of women who are more likely to identify as attempting or planning a pregnancy. These include economic status, age and relationship status. While these factors may typify women who are more likely to be proactively planning or attempting pregnancy, such women should not be seen to represent the only ones who warrant preconception care given the high rates of unplanned pregnancies among all women. In some regards, the differences between women who are currently attempting or planning pregnancy and other women reflect the areas of life that reproductive life planning advocates encourage women to consider before attempting pregnancy: namely, maternal health, age of the woman and her partner, and life context such as career development and marital status.^36^ As such, this finding should be seen as a call for policymakers to prioritise health promotion interventions that communicate the value of a reproductive life plan to the broader community, and the impacts of preconception health on pregnancy and child outcomes.^3^ However, it should also be acknowledged that there are a number of structural and political factors that may prevent some women from experiencing the control and self-determination required for a reproductive life plan^37^ and as such policymakers should also prioritise improving social and economic inequities in addition to providing individual family planning support to women. This policy work should be done in addition to campaigns and interventions that directly target the women already planning or attempting a pregnancy, with a focus on ensuring they have access to accurate and evidence-based preconception health information in all clinical encounters.

### Limitations

This study’s limitations are primarily associated with the self-reported nature of the survey data.

Self-reported data is vulnerable to recall or participant bias, particularly as responders were asked to answer several questions within the context of the previous 12 months. To some degree, this was counterbalanced by the inclusion of validated health service research instruments in the survey. The representative sample of the Australian population also offers some generalisability of the results. The analysis is also limited by the smaller sample sizes associated with the variables of interest which resulted in wide confidence intervals for some outcomes and limited power for further adjusted analysis meaning the results from Table 4 should be viewed as exploratory and interpreted cautiously.

## Conclusions

This manuscript presents an examination of the health service use of women planning or attempting pregnancy in Australia. The analysis found a wide range of health services are being used, including some health professions previously overlooked in Australia’s existing preconception health policy and planning. It also identified a range of sociodemographic characteristics associated with planning or attempting pregnancy that align with reproductive life planning advice. These findings suggest an important role for policymakers and health professionals to improve awareness of the importance of reproductive planning and preconception care in the wider community.

## Data Availability

Data is available from the authors upon reasonable request

## Acknowledgements

The authors would like to acknowledge Dr Erica McIntyre (Institute for Sustainable Futures, University of Technology Sydney) for her contribution to the design of the survey instrument.

## References

1. Stephenson J, Heslehurst N, Hall J, et al. Before the beginning: nutrition and lifestyle in the preconception period and its importance for future health. The Lancet. 2018;391(10132):1830–1841.

2. Caut C, Schoenaker D, McIntyre E, Vilcins D, Gavine A, Steel A. Relationships between women’s and men’s modifiable preconception risks and health behaviors and maternal and offspring health outcomes: an umbrella review. Semin Reprod Med. 2022;40(3-04):170–183.

3. Hall J, Chawla M, Watson D, et al. Addressing reproductive health needs across the life course: an integrated, community-based model combining contraception and preconception care. Lancet Public Health. Jan 2023;8(1):e76–e84. doi:10.1016/s2468-2667(22)00254-7

4. Barker M, Dombrowski SU, Colbourn T, et al. Intervention strategies to improve nutrition and health behaviours before conception. The Lancet. 2018;391(10132):1853–1864.

5. Daly M, Kipping RR, Tinner LE, Sanders J, White JW. Preconception exposures and adverse pregnancy, birth and postpartum outcomes: Umbrella review of systematic reviews. Paediatr Perinat Epidemiol. Mar 2022;36(2):288–299. doi:10.1111/ppe.12855

6. Withanage NN, Botfield JR, Srinivasan S, Black KI, Mazza D. Effectiveness of preconception interventions in primary care: a systematic review. Br J Gen Pract. Dec 2022;72(725):e865–e872. doi:10.3399/bjgp.2022.0040

7. Wald NJ. Folic acid and neural tube defects: Discovery, debate and the need for policy change. J Med Screen. Sep 2022;29(3):138–146. doi:10.1177/09691413221102321

8. Dorney E, Boyle J, Walker R, et al. A Systematic Review of Clinical Guidelines for Preconception Care. Semin Reprod Med. 2022;40(3-04):157–169.

9. Lang AY, Harrison CL, Barrett G, Hall JA, Moran LJ, Boyle JA. Opportunities for enhancing pregnancy planning and preconception health behaviours of Australian women. Women and Birth. 2021;34(2):e153–e161.

10. Ranatunga I, Jayaratne K. Proportion of unplanned pregnancies, their determinants and health outcomes of women delivering at a teaching hospital in Sri Lanka. BMC Pregnancy Childbirth. Nov 5 2020;20(1):667. doi:10.1186/s12884-020-03259-2

11. Wellings K, Jones KG, Mercer CH, et al. The prevalence of unplanned pregnancy and associated factors in Britain: findings from the third National Survey of Sexual Attitudes and Lifestyles (Natsal-3). Lancet. Nov 30 2013;382(9907):1807–16. doi:10.1016/s0140-6736(13)62071-1

12. Backhausen MG, Ekstrand M, Tydén T, et al. Pregnancy planning and lifestyle prior to conception and during early pregnancy among Danish women. Eur J Contracept Reprod Health Care. Feb 2014;19(1):57–65. doi:10.3109/13625187.2013.851183

13. Wellings K, Jones KG, Mercer CH, et al. The prevalence of unplanned pregnancy and associated factors in Britain: findings from the third National Survey of Sexual Attitudes and Lifestyles (Natsal-3). The Lancet. 2013;382(9907):1807–1816. doi:10.1016/s0140-6736(13)62071-114.

14. Daly MP, White J, Sanders J, Kipping RR. Women’s knowledge, attitudes and views of preconception health and intervention delivery methods: a cross-sectional survey. BMC Pregnancy Childbirth. Sep 24 2022;22(1):729. doi:10.1186/s12884-022-05058-3

15. McGowan L, Lennon-Caughey E, Chun C, McKinley MC, Woodside JV. Exploring preconception health beliefs amongst adults of childbearing age in the UK: a qualitative analysis. BMC Pregnancy Childbirth. Jan 16 2020;20(1):41. doi:10.1186/s12884-020-2733-5

16. Musgrave L, Homer C, Gordon A. Knowledge, attitudes and behaviours surrounding preconception and pregnancy health: an Australian cross-sectional survey. BMJ Open. Jan 3 2023;13(1):e065055. doi:10.1136/bmjopen-2022-065055

17. Schoenaker D, Stephenson J, Smith H, et al. Women’s preconception health in England: a report card based on cross-sectional analysis of national maternity services data from 2018/2019. Bjog. Feb 21 2023; doi:10.1111/1471-0528.17436

18. McDougall B, Kavanagh K, Stephenson J, Poston L, Flynn AC, White SL. Health behaviours in 131,182 UK women planning pregnancy. BMC Pregnancy Childbirth. Jul 28 2021;21(1):530. doi:10.1186/s12884-021-04007-w

19. Wise LA, Wesselink AK, Hatch EE, et al. Changes in Behavior with Increasing Pregnancy Attempt Time: A Prospective Cohort Study. Epidemiology. Sep 2020;31(5):659–667. doi:10.1097/ede.0000000000001220

20. Schoenaker D, Connolly A, Stephenson J. Preconception care in primary care: supporting patients to have healthier pregnancies and babies. Br J Gen Pract. Apr 2022;72(717):152. doi:10.3399/bjgp22X718853

21. World Health Organization. Women of reproductive age (15-49 years) population (thousands). World Health Organization. Accessed 9th November, 2022. https://www.who.int/data/gho/indicator-metadata-registry/imr-details/women-of-reproductive-age-(15-49-years)-population-(thousands)

22. Cummins R, Eckersley R, Pallant JF, Van Vugt JMG, Misajon R. Developing a national index of subjective wellbeing: The Australian Unity Wellbeing Index. Social Indicators Research. 2003;64(2):159–90.

23. Australian Centre on Quality of Life. Subjective wellbeing in Australia during the second year of the pandemic. 2022. https://www.acqol.com.au/publications#reports

24. Hays RD, Sherbourne CD, Mazel R. User’s Manual for the Medical Outcomes Study (MOS) Core Measures of Health-Related Quality of Life. 1995. MR-162-RC. https://www.rand.org/pubs/monograph_reports/MR162.html

25. Australian Institute of Health and Welfare. Primary health care in Australia. Australian Government. Accessed 10 July, 2023. https://www.aihw.gov.au/reports/primary-health-care/primary-health-care-in-australia/contents/about-primary-health-care

26. Walzman M. Every contact is an opportunity to promote healthy lifestyles. BMJ : British Medical Journal. 2014;349:g7222. doi:10.1136/bmj.g7222

27. Stephenson J, Patel D, Barrett G, et al. How do women prepare for pregnancy? Preconception experiences of women attending antenatal services and views of health professionals. PloS one. 2014;9(7):e103085.

28. Cha E, Smart MJ, Braxter BJ, Faulkner MS. Preconception care to reduce the risks of overweight and obesity in women of reproductive age: an integrative review. International Journal of Environmental Research and Public Health. 2021;18(9):4582.

29. Steel A, Adams J, Sibbritt D. The Characteristics of Women Who Use Complementary Medicine While Attempting to Conceive: Results from a Nationally Representative Sample of 13,224 Australian Women. Women’s Health Issues. 2017;27(1):67–74.

30. Xie Z-y, Peng Z-h, Yao B, et al. The effects of acupuncture on pregnancy outcomes of in vitro fertilization: a systematic review and meta-analysis. BMC Complementary and Alternative Medicine. 2019/06/14 2019;19(1):131. doi:10.1186/s12906-019-2523-7

31. Steel A, Adams J, Sibbritt D, Broom A, Gallois C, Frawley J. Determinants of women consulting with a complementary and alternative medicine practitioner for pregnancy-related health conditions. Women & health. 2014;54(2):127–144.

32. Fernandez M, Young A, Milton K, et al. Physical activity promotion in chiropractic: a systematic review of clinician-based surveys. Chiropractic & Manual Therapies. 2022;30(1):1–13.

33. Fikar PE, Edlund KA, Newell D. Current preventative and health promotional care offered to patients by chiropractors in the United Kingdom: a survey. Chiropractic & manual therapies. 2015;23:1–7.

34. Pinto JW, Bradbury K, Newell D, Bishop FL. Lifestyle and health behavior change in traditional acupuncture practice: a systematic critical interpretive synthesis. The Journal of Alternative and Complementary Medicine. 2021;27(3):238–254.

35. Pinto J, Bradbury K, Newell D, Bishop F. Lifestyle and health behaviour change support in traditional acupuncture: a mixed method survey study of reported practice (UK). BMC Complementary Medicine and Therapies. 2022;22(1):248.

36. Files JA, Frey KA, David PS, Hunt KS, Noble BN, Mayer AP. Developing a Reproductive Life Plan. https://doi.org/10.1111/j.1542-2011.2011.00048.x. Journal of Midwifery & Women’s Health. x2011/09/01 2011;56(5):468–474. doi:10.1111/j.1542-2011.2011.00048.x

37. Ending the postcode lottery: addressing barriers to sexual, maternity and reproductive healthcare in Australia (2023).

